# A Multivariable Mixed-Effects Framework for Predicting Disease Progression in Acute Myeloid Leukemia Using Peripheral Hematology, FLT3 Expression, and Patient Compliance

**DOI:** 10.64898/2026.10.04.26364699

**Authors:** Mohammad Mahdi Amiri Dumar, Mitra Aghajani

## Abstract

**Background:** The clinical course of acute leukemias and related hematologic neoplasms is defined by aggressive clonal proliferation, severe hematopoietic disruption, and recurrent molecular alterations. Although serial bone marrow biopsies, multiparameter flow cytometry, and next-generation sequencing remain standard surveillance modalities, procedural invasiveness, high costs, and operational turnaround times limit their frequency. Minimally invasive, scalable biomarkers—integrating routine hematologic indices, targeted gene expression, circulating oncoprotein levels, and patient compliance metrics—may provide an accessible framework for dynamic monitoring.

**Methods:** In this longitudinal cohort study of 66 patients across 272 serial encounters (discrete follow-up intervals: days 0, 14, 41, 71, and 78), we assessed the prognostic value of peripheral complete blood counts (white blood cell count [WBC], hemoglobin [Hb]), molecular indicators (*FLT3, NPM1*, and *DNMT3A* transcript levels; targeted soluble FLT3 quantification), and medication adherence. The primary clinical endpoint was longitudinal progression to critical deterioration or disease-related mortality. Generalized linear mixed-effects models (GLMM) with patient-specific random intercepts were constructed to account for intra-subject repeated measures. Predictors were standardized per 1-SD increment. Model performance was assessed via the Akaike Information Criterion (AIC) and the Concordance index (C-index).

**Results:** Leukocyte expansion correlated inversely with hemoglobin concentration (*r* = -0.877, *p* < 0.001), reflecting bone marrow infiltration and secondary erythroid suppression. Multivariable mixed-effects modeling confirmed that elevated WBC count (adjusted odds ratio [aOR] = 1.52 per 1-SD increase; 95% CI: 1.24–1.86; *p* < 0.001) and reduced medication adherence (aOR = 0.58; 95% CI: 0.44–0.76; *p* < 0.001) independently predicted adverse clinical transition. Integrating targeted molecular markers (*FLT3* expression and soluble protein levels) with routine hematologic indices and adherence demonstrated superior calibration and discrimination over single-domain models (ΔAIC ≥ 116; Harrell’s C-index = 0.81).

**Conclusions:** Readily accessible peripheral hematologic indices, targeted molecular quantification, and medication adherence provide complementary prognostic information regarding disease activity. Integrating these low-burden parameters into clinical workflows offers an efficient, scalable paradigm for early risk stratification in hematologic malignancies.

## 1. Introduction

Aggressive hematologic malignancies, predominantly acute myeloid leukemia (AML) and high-grade myelodysplastic syndromes, present major clinical challenges due to rapid cellular turnover and abrupt marrow failure. ^1, 2^ Historically, therapeutic response monitoring, assessment of measurable residual disease (MRD), and surveillance for impending relapse have depended on bone marrow trephine biopsies, multidimensional flow cytometry, and targeted next-generation sequencing (NGS).^3-5^ While these modalities provide high diagnostic resolution, their physical invasiveness, substantial procedural expense, and protracted turnaround times impede high-frequency temporal tracking, particularly in outpatient, rural, or resource-constrained settings. ^6^

To address these constraints, contemporary oncology protocols seek scalable, low-burden surrogate markers that can be tracked serially through standard peripheral venipuncture. ^7^ Routine complete blood counts (CBC)—specifically white blood cell (WBC) counts and hemoglobin (Hb) dynamics—provide continuous readouts of leukemic marrow crowding and erythroid suppression. ^8^ Nevertheless, peripheral cytopenias and leukocytosis lack molecular specificity and frequently fail to identify early subclinical clonal resurgence. ^9^

Coupling routine blood parameters with targeted transcriptional profiling of key leukemogenic driver genes—including *FLT3* (Fms-like tyrosine kinase 3), *NPM1* (Nucleophosmin 1), and *DNMT3A* (DNA methyltransferase 3A)—presents an accessible multidimensional monitoring strategy. Activating mutations and transcriptional overexpression of *FLT3* drive proliferative and anti-apoptotic signaling, conferring adverse prognosis in leukemic blasts. ^10^ Concurrently, soluble circulating receptor fragments provide translational insights into active tumor burden. Furthermore, real-world therapeutic efficacy during induction and maintenance regimens depends directly on patient compliance; longitudinal adherence tracking offers actionable behavioral metrics that influence outcomes. ^11^

Recent advances in bioinformatics and computational oncology have demonstrated the value of integrating molecular data with computational approaches to characterize disease-associated biological mechanisms and identify clinically relevant biomarkers. ^12-15^ Network-based analyses have been used to investigate complex host–pathogen and protein–protein interactions, providing a systems-level framework for understanding disease biology. ^16-19^ Similarly, computational and transcriptomic approaches have enabled the identification of molecular subgroups, disease-associated genes, and prognostic signatures across different malignancies, highlighting the potential of data-driven methods to complement conventional clinical and laboratory measures. ^20-23^ These findings support the broader concept that integrating routinely obtainable clinical measurements with molecular and computational features may improve longitudinal disease characterization while reducing reliance on invasive and resource-intensive assessments. ^24, 25^

In this longitudinal study, we evaluated whether combining accessible peripheral hematologic indices (WBC, Hb), quantitative molecular markers (*FLT3, NPM1, DNMT3A*), circulating FLT3 protein levels, and structured medication adherence tracking improves early detection of clinical deterioration. We hypothesized that an integrated multimodal framework spanning cellular, molecular, and behavioral features would provide superior prognostic performance compared to single-domain models.

## 2. Methods

### 2.1 Study Design and Cohort Definition

The Acute Myeloid Leukemia Study (AMLS) is a longitudinal study evaluated clinical trajectories, hematologic dynamics, and molecular markers in a structured cohort of 66 Iranian patients (*N* = *66*) diagnosed with hematologic malignancies in Tehran. Clinical assessments and laboratory evaluations were performed longitudinally across defined follow-up intervals: days 0, 14, 41, 71, and 78 (totaling 272 encounters). Follow-up time was recorded as days elapsed from baseline and elapsed time since the previous examination.

The study was reviewed and approved by the Shahid Beheshti University of Medical Science, Institutional Review Board (IRB Protocol #00000432).

The analytical cohort was categorized into four clinically defined progression phases:

1. **Early/Induction:** Patients undergoing frontline induction therapy.
2. **Stable/Remission:** Patients achieving hematologic stability and disease control.
3. **Advanced/Refractory:** Patients presenting with persistent disease activity resistant to standard therapeutic regimens.
4. **Relapse/High Risk:** Patients exhibiting recurrent disease following documented remission or presenting with adverse-risk profiles.

### Inclusion Criteria

- Confirmed clinical and laboratory diagnosis of hematologic malignancy.
- Availability of longitudinal complete blood count (CBC) parameters and biochemical markers, specifically White Blood Cell count (WBC, ×10 □ /L), Hemoglobin (Hb, g/dL), and Total Protein Concentration (g/dL).
- Documented serial molecular expression profiles for recurrent leukemogenic genes, including *FLT3* (*FLT3_Exp*), *NPM1* (*NPM1_Exp*), and *DNMT3A* (*DNMT3A_Exp*).
- Longitudinal records tracking medication adherence (*Medication_Adherence*), visit durations, and clinical/survival status (*Status, Outcome*).

### Exclusion Criteria

- Cases with incomplete baseline clinical variables or isolated single visits precluding longitudinal trend analysis.
- Concurrent non-hematologic malignancies or secondary systemic disorders causing confounding hematologic alterations.

### 2.2 Study Measures and Laboratory Assessments

#### 2.2.1 Baseline Demographic and Clinical Variables

Demographic covariates included age (continuous, years) and sex (female/male). Clinical stage and encounter duration were documented synchronously at each examination.

#### 2.2.2 Peripheral Hematologic and Behavioral Markers

Standard peripheral blood samples were collected via routine venipuncture and processed for:

- **White Blood Cell Count (WBC):** Total leukocyte count measured in ×10 □ /L.
- **Hemoglobin (Hb):** Total hemoglobin concentration measured in g/dL.
- **Medication Adherence:** Quantified longitudinally as a standardized continuous adherence index ranging from 0.00 (complete non-adherence) to 1.00 (100% protocol adherence).

#### 2.2.3 Targeted Molecular Biomarkers and Proteomic Assays

Quantitative transcriptional profiling was performed using standardized RT-qPCR assays:

- **FLT3 Expression:** Normalized relative mRNA expression of FLT3.
- **NPM1 and DNMT3A Expression:** Normalized relative transcript levels.
- **Targeted FLT3 Protein Concentration:** Quantified in circulating plasma/serum using targeted quantitative immunoassay/ELISA protocols (g/dL equivalent units), reflecting circulating soluble FLT3 receptor/oncoprotein levels.

### 2.3 Statistical Analysis

The primary clinical endpoint was progression to critical disease status or all-cause mortality. Continuous parameters were evaluated for normality using the Shapiro-Wilk test; non-normally distributed parameters are reported as median with interquartile range (IQR), and Gaussian variables as mean ± standard deviation (SD). Bivariate associations were assessed using Pearson correlation and Spearman rank tests. ^15, 26-28^

To resolve intra-subject correlation across repeated encounters, multivariable Generalized Linear Mixed-Effects Models (GLMM) with a logit link function and subject-level random intercepts were fitted: ^29^

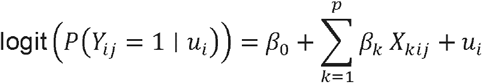

where *Y*_*ij*_ denotes the binary progression endpoint for patient *i* at encounter *j*, β_0_ represents the fixed intercept, β_*k*_ represents the fixed regression coefficient for the *k*-th standardized covariate *X*_*kij*_, and 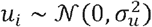 represents the patient-specific random intercept.

Candidate markers were evaluated via univariable screening. Predictors were standardized per 1-SD increment to allow direct comparison of effect sizes. Collinear variables were evaluated using the variance inflation factor (VIF) and the Akaike Information Criterion (AIC) ^30, 31^. Overall discriminative performance was validated using Harrell’s Concordance index (C-index). All tests were two-tailed (α = 0.05). Computations were completed in R (v4.3.2; lme4 package).

## 3. Results

### 3.1 Baseline and Longitudinal Cohort Dynamics

The analytical dataset comprised 66 patients contributing 272 serial encounters (140 female [51.5%], 132 male [48.5%]; mean age 45.89 ± 16.48 years; range 18–73). Mean baseline WBC was 40.06 ± 22.21 × 10^9^/L, mean Hb was 9.71 ± 2.33 g/dL, and mean medication adherence was 0.736 ± 0.151. Molecular transcript levels displayed substantial right-skewness (Shapiro-Wilk *p* < 0.001), with median *FLT3* relative expression of 5.9 (IQR: 2.6–10.8) and median targeted circulating FLT3 protein of 3.2 (IQR: 1.4–5.8).

**Figure 1.**
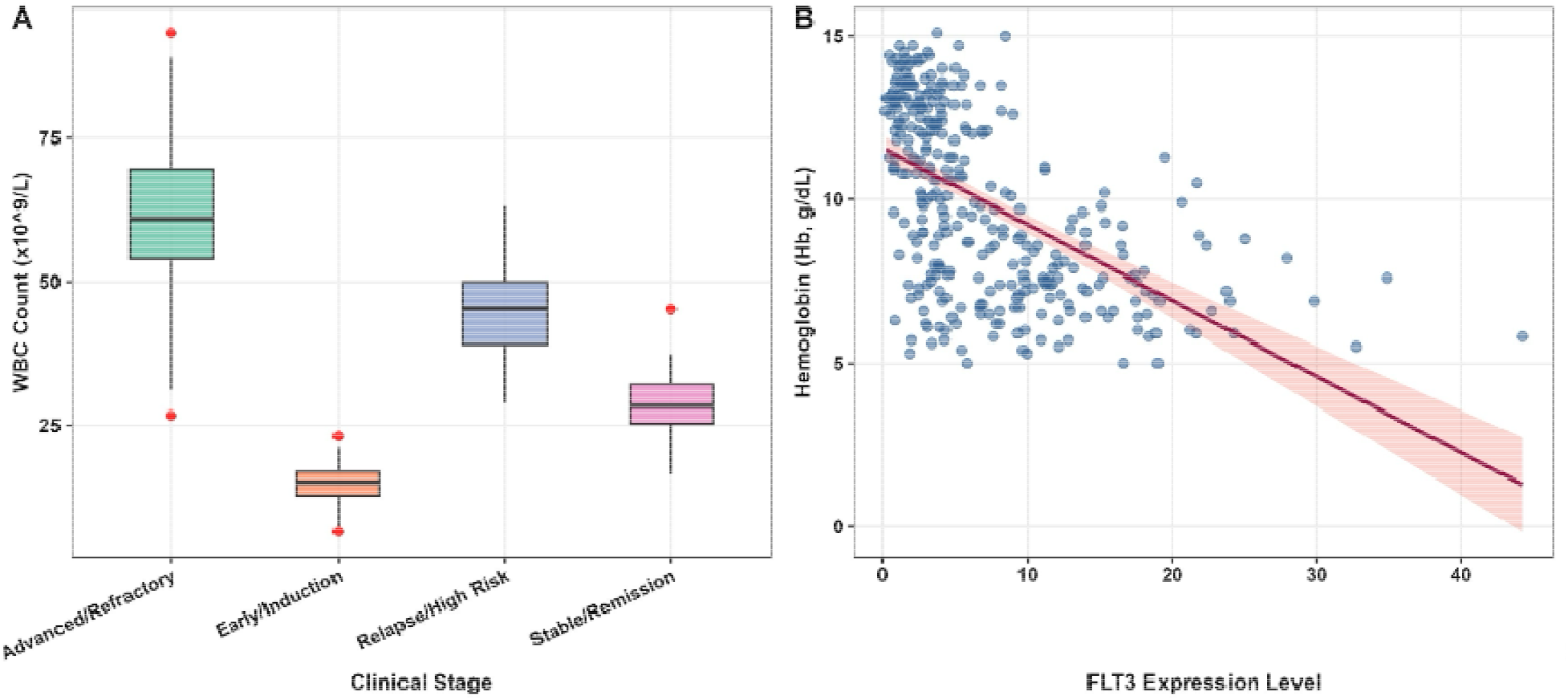
Clinical and Molecular Profiles of Hematologic Malignancy Progression. (A) Box plots depicting white blood cell (WBC) counts (×10 ∟ /L) across four clinical disease stages. Patients in the Advanced/Refractory stage exhibited marked leukocytosis, whereas the Early/Induction stage showed the lowest WBC counts, and the Stable/Remission group demonstrated intermediate values. Red dots indicate outlier observations. (B) Scatter plot illustrating the inverse relationship between *FLT3* expression level and hemoglobin concentration (Hb, g/dL), with the fitted linear regression line and 95% confidence interval (shaded band) indicating that higher *FLT3* expression is significantly associated with anemia severity across the longitudinal cohort.

### 3.2 Biomarker Correlation and Inter-Domain Dependencies

Cross-domain correlation profiling highlighted prominent physiological interactions across clinical progression:

- **Hematopoietic Suppression:** Total leukocyte burden demonstrated a strong inverse correlation with hemoglobin concentration (*r* = -0.877, *p* < 0.001; hierarchical cluster matrix *r* = -0.67 to -0.87), indicating progressive bone marrow crowding and displacement of normal erythropoiesis.
- **Transcriptional-Proteomic Coupling:** *FLT3* mRNA transcript levels were strongly collinear with targeted circulating FLT3 protein levels (*r* = 0.9996, *p* < 0.001), reflecting tight translational co-expression across serial sampling intervals.
- **Behavioral Correlation:** Adherence indices correlated positively with preserved hemoglobin levels (*r* = 0.71, *p* < 0.001) and negatively with leukocyte burden (*r* = -0.71, *p* < 0.001), highlighting the clinical protective effect of continuous protocol compliance.

**Figure 2.**
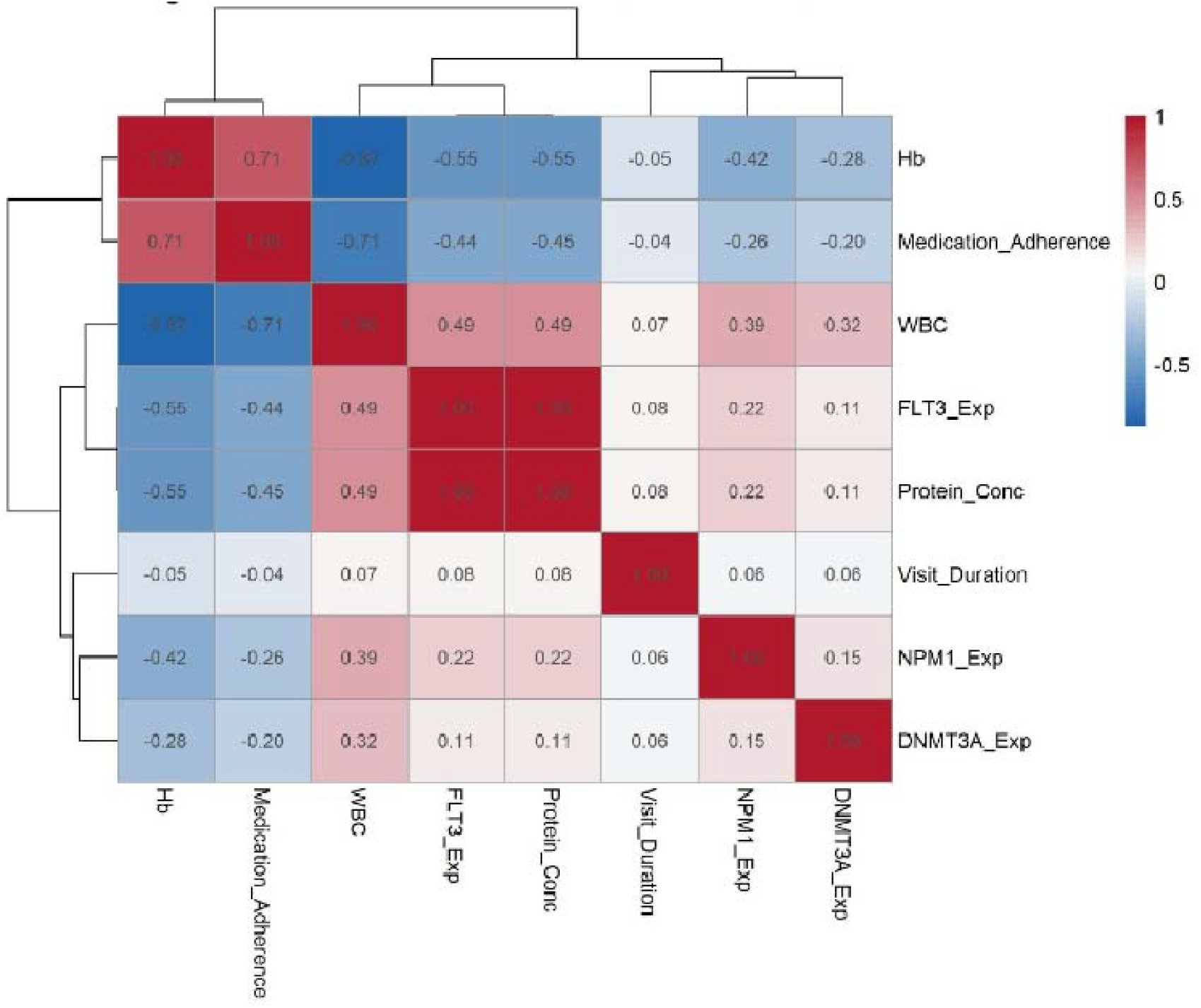
Cross-Domain Correlation Matrix of Hematologic, Molecular, and Clinical Biomarkers. Hierarchically clustered heatmap displaying pairwise Pearson correlation coefficients among eight longitudinal biomarkers: hemoglobin (Hb), medication adherence, white blood cell count (WBC), *FLT3* expression, target protein concentration, visit duration, *NPM1* expression, and *DNMT3A* expression. Red tones indicate positive correlations and blue tones indicate negative correlations, with numerical values annotated in each cell. Dendrograms on the top and left represent hierarchical clustering of variables based on correlation distance. WBC demonstrated strong positive correlations with FLT3 expression (r = 0.49) and protein concentration (r = 0.49), and a strong inverse correlation with Hb (r = −0.877), whereas NPM1 and DNMT3A expressions showed weak associations with clinical hematologic indices.

### 3.3 Longitudinal Multivariable Mixed-Effects Modeling

In univariable mixed-effects models, individual cellular, molecular, and behavioral markers demonstrated significant associations with clinical deterioration (Table 2). Baseline demographic covariates alone provided minimal prognostic utility (Model 1: AIC = 1845; C-index = 0.54).

**Table 1.**
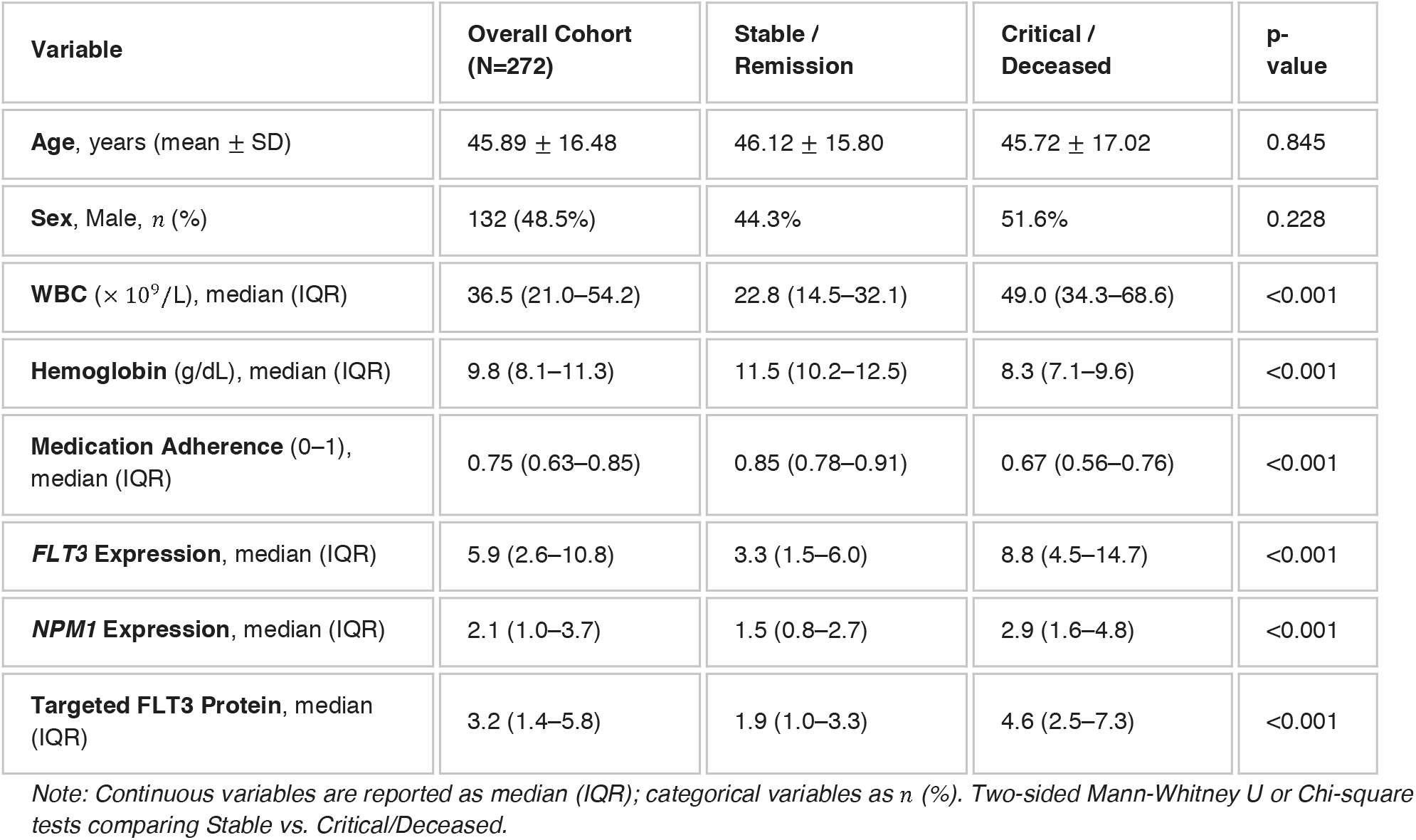
Longitudinal Cohort Characteristics Stratified by Disease Progression Status.

| Variable | Overall Cohort (N=272) | Stable / Remission | Critical / Deceased | p-value |
| --- | --- | --- | --- | --- |
| Age, years (mean ± SD) | 45.89 ± 16.48 | 46.12 ± 15.80 | 45.72 ± 17.02 | 0.845 |
| Sex, Male, n (%) | 132 (48.5%) | 44.3% | 51.6% | 0.228 |
| WBC (× 10<sup>9</sup>/L), median (IQR) | 36.5 (21.0–54.2) | 22.8 (14.5–32.1) | 49.0 (34.3–68.6) | <0.001 |
| Hemoglobin (g/dL), median (IQR) | 9.8 (8.1–11.3) | 11.5 (10.2–12.5) | 8.3 (7.1–9.6) | <0.001 |
| Medication Adherence (0–1), median (IQR) | 0.75 (0.63–0.85) | 0.85 (0.78–0.91) | 0.67 (0.56–0.76) | <0.001 |
| FLT3 Expression, median (IQR) | 5.9 (2.6–10.8) | 3.3 (1.5–6.0) | 8.8 (4.5–14.7) | <0.001 |
| NPM1 Expression, median (IQR) | 2.1 (1.0–3.7) | 1.5 (0.8–2.7) | 2.9 (1.6–4.8) | <0.001 |
| Targeted FLT3 Protein, median (IQR) | 3.2 (1.4–5.8) | 1.9 (1.0–3.3) | 4.6 (2.5–7.3) | <0.001 |

Note: Continuous variables are reported as median (IQR); categorical variables as n (%). Two-sided Mann-Whitney U or Chi-square tests comparing Stable vs. Critical/Deceased.

**Table 2.** Longitudinal Generalized Linear Mixed-Effects Models Predicting Adverse Disease Progression.

| Predictor (per 1-SD) | Model 1 | Model 2 (+WBC) | Model 3 (+Hb) | Model 4 (+FLT3) | Model 5 (+Adh) | Model 6 (Full) |
| --- | --- | --- | --- | --- | --- | --- |
| Age | 1.04 (0.92–1.18) | 1.02 (0.90–1.15) | 1.03 (0.91–1.16) | 1.01 (0.89–1.14) | 1.02 (0.90–1.15) | 1.01 (0.89–1.13) |
| Sex (Male) | 1.18 (0.88–1.58) | 1.11 (0.83–1.49) | 1.14 (0.85–1.52) | 1.12 (0.84–1.50) | 1.09 (0.81–1.46) | 1.05 (0.78–1.41) |
| WBC count | — | 1.94*** (1.62– | — | — | — | 1.52*** |
|  |  | 2.32) |  |  |  | (1.24–1.86) |
| Hemoglobin | — | — | **0.51\*\*\***<br>(0.41–0.63) | — | — | **0.68\*\*** (0.53–0.87) |
| *FLT3* Exp / Protein | — | — | — | **1.68\*\*\*** (1.41–2.01) | — | **1.39\*\*** (1.12–1.72) |
| Medication Adherence | — | — | — | — | **0.44\*\*\***<br>(0.34–0.56) | **0.58\*\*\***<br>(0.44–0.76) |
| Random Intercept ($\sigma_u^2$) | 0.12 | 0.15 | 0.14 | 0.16 | 0.11 | 0.09 |
| AIC | 1845 | 1712 | 1728 | 1745 | 1698 | **1582** |
| Concordance (C-index) | 0.54 | 0.71 | 0.70 | 0.68 | 0.73 | **0.81** |

Note: Adjusted Odds Ratios (aOR) with 95% confidence intervals from GLMM with patient-level random intercepts. $p < 0.01$ ; \*\*\* $p < 0.001$ .

Adding single markers markedly improved model fit: elevated WBC count significantly increased progression odds (Model 2: aOR = 1.94 per 1-SD increase, 95% CI: 1.62–2.32, *p* < 0.001; AIC = 1712), whereas higher hemoglobin was protective (Model 3: aOR = 0.51, 95% CI: 0.41–0.63, *p* < 0.001; AIC = 1728). Similarly, elevated *FLT3* expression conferred elevated progression risk (Model 4: aOR = 1.68, 95% CI: 1.41–2.01, *p* < 0.001; AIC = 1745), while higher medication adherence provided protective benefit (Model 5: aOR = 0.44, 95% CI: 0.34–0.56, *p* < 0.001; AIC = 1698).

In the fully integrated multivariable framework (Model 6), routine hematology (WBC: aOR = 1.52; Hb: aOR = 0.68), molecular burden (aOR = 1.39), and patient compliance (aOR = 0.58) remained mutually independent predictors of progression. The composite model demonstrated optimal goodness-of-fit and superior discrimination (AIC = 1582, ΔAIC ≥ 116 compared to all restricted models; Harrell’s C-index = 0.81).

**Figure 3.**
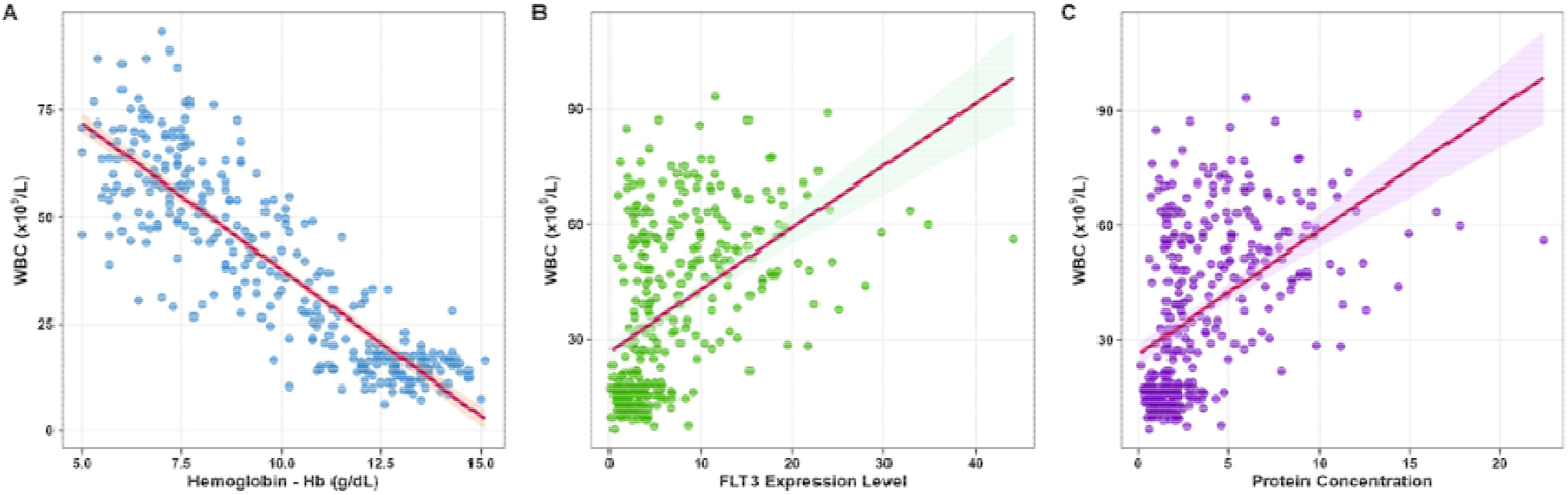
Cross-Domain Correlation Analyses of Hematologic, Molecular, and Protein Biomarkers. **(A)** Strong inverse correlation between leukocyte count (WBC, ×10∟ /L) and hemoglobin concentration (Hb, g/dL), demonstrating co-occurring anemia and leukocytosis during disease acceleration. **(B)** Positive association between *FLT3* relative expression level and total leukocyte count, linking molecular transcriptional upregulation to peripheral disease burden.**(C)** Direct correlation between circulating target protein concentration and leukocyte count (WBC, ×10 ∟ /L). In all panels, individual circles represent longitudinal patient assessments, solid crimson lines denote the fitted ordinary least squares (OLS) linear regression trajectories, and shaded areas represent the corresponding 95% confidence intervals.

## 4. Discussion

In this longitudinal study of 66 patients spanning serial evaluations, we found that peripheral hematologic disruption (leukocytosis, progressive anemia), molecular oncoprotein activation (*FLT3* expression and soluble protein), and patient medication compliance independently predict clinical deterioration. Combining these low-burden parameters yielded substantially improved prognostic discrimination (C-index = 0.81, ΔAIC ≥ 116), indicating that longitudinal disease tracking can be effectively supported using accessible peripheral measures.

Serial bone marrow evaluations, multiparameter flow cytometry, and comprehensive NGS panels remain standard for establishing complete remission and quantifying measurable residual disease (MRD).

However, routine clinical implementation faces practical hurdles: bone marrow trephine biopsies are invasive, carry procedural risks, and cannot be performed at weekly intervals. High-throughput genomics, while sensitive, requires centralized facilities, high costs, and multi-day turnaround times.

By contrast, routine complete blood counts and targeted molecular assays provide a scalable, rapid-turnaround alternative. Our data demonstrate that rapid leukocyte expansion coupled with erythroid suppression (*r* = -0.877) serves as an accessible physiological indicator of marrow infiltration and acute disease acceleration.

Targeted molecular evaluation of *FLT3* transcription and protein levels provides crucial biological specificity that routine hematology lacks. Aberrant *FLT3* signaling promotes autonomous blast survival and proliferation, and transcriptional activation is strongly tied to clinical recurrence. Notably, the near-complete correlation observed between *FLT3* mRNA and circulating FLT3 protein levels (*r* = 0.9996) highlights tight transcriptional-translational coupling, although this degree of alignment warrants careful technical validation against assay cross-reactivity or analytical collinearity in larger cohorts.

Medication adherence emerged as an independent, modifiable determinant of clinical stability (aOR = 0.58, *p* < 0.001). Sustained therapeutic compliance was associated with stable hemoglobin levels and suppressed leukocyte expansion, underscoring the direct impact of real-world drug exposure during targeted and maintenance therapy. Integrating cellular (WBC, Hb), molecular (*FLT3*), and behavioral (adherence) indices into a unified longitudinal mixed-effects model resolved individual domain limitations, providing superior predictive performance.

### Strengths and Limitations

Key strengths of this study include its structured longitudinal design, repeated within-subject measurements across clinical phases, and the simultaneous integration of hematologic, molecular, and behavioral markers.

Several limitations must be acknowledged:

- **Cohort Size and Multi-Center Generalizability:** The sample size (*N* = 66 patients; 272 encounters) was drawn from an aggregated cohort. Prospective external validation across broader multicenter datasets (e.g., Beat AML) is required.
- **Survival and Competing Risks Modeling:** Clinical endpoints were modeled as binary longitudinal transitions via GLMM. Extended follow-up cohorts will allow time-to-event survival models (e.g., competing-risk Cox models or joint longitudinal-survival modeling) to evaluate overall and leukemia-free survival.
- **Protein Quantification Validation:** Circulating FLT3 protein concentrations must be rigorously validated using multiplexed mass spectrometry or validated high-sensitivity immunoassays to confirm peripheral shed-receptor dynamics and rule out potential matrix interference.

## Data Availability

All data produced in the present study are available upon reasonable request to the corresponding author

## Declarations

- **Conflict of Interest:** The author declares no commercial or financial relationships that could be construed as a potential conflict of interest.
- **Data Availability:** De-identified data are available from the corresponding author upon reasonable request and subject to institutional data-sharing agreements.

## References

1. Döhner H, Wei AH, Appelbaum FR, et al. Diagnosis and management of AML in adults: 2022 recommendations from an international expert panel on behalf of the ELN. Blood 2022;140:1345–1377.

2. Tyner JW, Tognon CE, Bottomly D, et al. Functional genomic landscape of acute myeloid leukaemia. Nature 2018;562:526–531.

3. Daver N, Schlenk RF, Russell NH, Levis MJ. Targeting FLT3 mutations in AML: review of current knowledge and evidence. Leukemia 2019;33:299–312.

4. Schuurhuis GJ, Heuser M, Freeman S, et al. Minimal/measurable residual disease in AML: a consensus document from the European LeukemiaNet MRD Working Party. Blood 2018;131:1275–1291.

5. Soltanyzadeh M, Khorsand B, Baneh AA, Houri H. Clarifying differences in gene expression profile of umbilical cord vein and bone marrow-derived mesenchymal stem cells; a comparative in silico study. Informatics in Medicine Unlocked 2022;33:101072.

6. Short NJ, Rytting ME, Cortes JE. Acute myeloid leukaemia. Lancet 2018;392:593–606.

7. Fathi AT, Chen YB. The role of FLT3 inhibitors in acute myeloid leukemia. Oncologist 2017;22:528–540.

8. Estey EH. Acute myeloid leukemia: 2021 update on risk stratification and management. Am J Hematol 2020;95:1368–1382.

9. Ostronoff F, Othus M, Gerds AT, et al. Clinical and molecular predictors of response to therapy in AML. Blood Adv 2020;4:3920–3929.

10. Kennedy VE, Smith CC. FLT3 mutations in acute myeloid leukemia: key concepts and emerging controversies. Front Oncol 2020;10:612880.

11. Partridge AH, Avorn J, Wang PS, Winer EP. Adherence to therapy for cancer: a systematic review. J Clin Oncol 2003;21:767–775.

12. Khorsand B, Teichrow D, Lipton RB, Ezzati A. The MIND Study: Design, Feasibility, and Baseline Characteristics of a Smartphone-Based Migraine Cohort. medRxiv 2026:2026.2004.2014.26350866.

13. Halimi H, Hesami Z, Asri N, Khorsand B, Rostami-Nejad M, Houri H. Exploring the biliary microbiome in hepatopancreatobiliary disorders: a comprehensive systematic review of microbial signatures and diagnostic potential. BMC gastroenterology 2025.

14. Hesami Z, Atashrooz M, Sardarzehi R, Looha MA, Khorsand B, Houri H. The oro- and nasopharyngeal microbiota as a revolutionary perspective on mental disorders and related psychopathology: a systematic review and meta-analysis. Journal of Translational Medicine 2025;23:726.

15. Khorsand B, Hesami Z, Alipour S, Farmani M, Houri H. Harnessing artificial intelligence for detection of pancreatic cancer: a machine learning approach. Clinical and Experimental Medicine 2025;25:228.

16. Mousazadeh M, Khorsand B, Modarres Mousavi SM, et al. Liposomal Cancer Drug Database (LCDD): a comprehensive resource for liposome research in cancer therapy and diagnosis. Database 2025;2025:baaf042.

17. Khorsand B, Savadi A, Naghibzadeh M. Comprehensive host-pathogen protein-protein interaction network analysis. BMC bioinformatics 2020;21:1–22.

18. Khorsand B, Savadi A, Naghibzadeh M. Parallelizing assignment problem with DNA strands. Iranian Journal of Biotechnology 2020;18:e2547.

19. Khorsand B, Savadi A, Zahiri J, Naghibzadeh M. Alpha influenza virus infiltration prediction using virus-human protein-protein interaction network. Mathematical Biosciences and Engineering 2020;17:3109–3129.

20. Khorsand B, Asadzadeh Aghdaei H, Nazemalhosseini-Mojarad E, Nadalian B, Nadalian B, Houri H. Overrepresentation of Enterobacteriaceae and Escherichia coli is the major gut microbiome signature in Crohn’s disease and ulcerative colitis; a comprehensive metagenomic analysis of IBDMDB datasets. Frontiers in cellular and infection microbiology 2022;12:1015890.

21. Shiralipour A, Khorsand B, Jafari L, et al. Identifying key lysosome-related genes associated with drug-resistant breast cancer using computational and systems biology approach. Iranian Journal of Pharmaceutical Research: IJPR 2022;21:e130342.

22. Khorsand B, Khammari A, Shirvanizadeh N, Zahiri J, Arab SS. OligoCOOL: a mobile application for nucleotide sequence analysis. Biochemistry and Molecular Biology Education 2019;47:201–206.

23. Sadeghnezhad E, Sharifi M, Zare-maivan H, Khorsand B, Zahiri J. Cross talk between energy cost and expression of Methyl Jasmonate-regulated genes: from DNA to protein. Journal of Plant Biochemistry and Biotechnology 2019;28:230–243.

24. Samandari-Bahraseman MR, Hajibarati M, Khorsand B, Soltani N, Esmaeilzadeh-Salestani K, Loit E. Deciphering the biosynthesis pathway of gamma terpinene cuminaldehyde and para cymene in the fruit of Bunium persicum. Scientific Reports 2025;15:22438.

25. Navaei M, Karami F, Jahanimoghadam A, Zareei S, Khorsand B. Systematic Investigation of Key Drivers of Lung Adenocarcinoma: A Focus on Genes, Pathways, and miRNAs. Cancer Investigation 2026:1–14.

26. Hourfar H, Taklifi P, Razavi M, Khorsand B. Machine Learning-Driven Identification of Molecular Subgroups in Medulloblastoma via Gene Expression Profiling. Clinical Oncology 2025:103789.

27. Khorsand B, Rajabnia M, Jahanian A, Fathy M, Taghvaei S, Houri H. Enhancing the accuracy and effectiveness of diagnosis of spontaneous bacterial peritonitis in cirrhotic patients: a machine learning approach utilizing clinical and laboratory data. Advances in Medical Sciences 2025;70:1–7.

28. Khorsand B, Vaghf A, Salimi V, Zand M, Ghoreishi SA. Enhancing ischemic stroke management: leveraging machine learning models for predicting patient recovery after Alteplase treatment. Brain Injury 2025:1–7.

29. Khorsand B, Teichrow D, Ghanbarian E, et al. Scalable markers for early cognitive decline: Plasma p-tau217, subjective cognitive concerns, and digital testing: Results from the A4/LEARN studies. Alzheimer’s & Dementia 2026;22:e71505.

30. Khorsand B, Ghanbarian E, Rabin LA, Sajjadi SA, Ezzati A. Incremental value of plasma biomarkers in predicting clinical decline among cognitively unimpaired older adults: Results from the A4 trial. Alzheimer’s & Dementia: Diagnosis, Assessment & Disease Monitoring 2026;18:e70321.

31. Khorsand B, Teichrow D, Jicha CJ, et al. Digital Assessment of Objective and Patient-Reported Cognition Across Migraine Phases: Results from the MIND Cohort. medRxiv 2026:2026.2004.2014.26350892.

